# Abnormal Lipid Profiles as Markers of Diabetic Macular Edema Among Patients with Type 2 Diabetes Mellitus Attending a Tertiary Hospital in Northern Tanzania: A One-Year Cross-Sectional Study

**DOI:** 10.64898/2026.03.03.26347512

**Authors:** Muhidini Huud Swalehe, William Makupa, Andrew Makupa, Rosina Deocar, Frank Sandi

## Abstract

**Background:** Diabetes mellitus (DM) remains a major global health challenge and is associated with vision-threatening complications, including diabetic macular edema (DME), a leading cause of visual impairment. Dyslipidemia has been implicated in the development of macular edema through mechanisms involving vascular permeability, endothelial dysfunction, and chronic inflammation. However, evidence regarding the relationship between lipid abnormalities and macular edema remains inconsistent across studies.

**Aim:** This study aimed to evaluate the association between abnormal lipid profiles and diabetic macular edema among patients with type 2 diabetes mellitus attending Kilimanjaro Christian Medical Centre (KCMC).

**Methods:** A hospital-based analytical cross-sectional study was conducted among 296 diabetic outpatients at KCMC. Participants underwent comprehensive ophthalmic evaluation including fundoscopy and imaging with optical coherence tomography (OCT) for assessment of macular edema. Blood samples were collected for biochemical lipid analysis. Data were cleaned and analyzed using STATA version 17.

**Results:** Diabetic macular edema was identified in 56.4% (167/296) of participants. Abnormal lipid parameters were common, with elevated total cholesterol observed in 48.6%, triglycerides in 43.6%, low-density lipoprotein (LDL) in 36.1%, and reduced high-density lipoprotein (HDL) in 38.9% of patients. Elevated total cholesterol, triglycerides, and LDL levels showed significant associations with macular edema (p < 0.05). After multivariable adjustment, serum triglycerides remained independently associated with macular edema (p = 0.002).

**Conclusion:** Dyslipidemia demonstrated a significant association with diabetic macular edema, with serum triglycerides emerging as an independent predictor. These findings highlight the importance of lipid monitoring, lifestyle modification, and strengthened screening strategies in reducing the burden of vision-threatening diabetic complications.

## 1. INTRODUCTION

Diabetes mellitus (DM) is a metabolic disorder characterized by chronic hyperglycemia resulting from impaired insulin secretion, insulin action, or both. Lifestyle factors such as obesity, reduced physical activity, and sedentary behavior significantly contribute to the increasing burden of diabetes worldwide (Sajjan and Shamsuddin, 2016). It can either be type 1 diabetes that occurs due to absolute insulin deficiency, or type 2 diabetes results from insulin resistance and relative insulin insufficiency (WHO).

Persistent hyperglycemia and metabolic disturbances in DM lead to progressive microvascular damage affecting multiple organ systems, including the eyes, kidneys, vascular system, and nervous system (Pandey, 2021). The global prevalence of diabetes has increased markedly, rising from approximately 30 million cases in 1985 to 177 million in 2000 (Rasoulinejad et al., 2015), with projections exceeding 360 million affected individuals by 2030 (S, S.S and M.K., 2021).

Ocular complications such as diabetic retinopathy (DR) and diabetic macular edema (DME) remain leading causes of visual impairment worldwide (Rasoulinejad et al., 2015). Vision loss in diabetes is primarily attributed to retinal microvascular damage, capillary leakage causing macular edema, and macular ischemia secondary to capillary occlusion (Rasoulinejad et al., 2015; Pandey, 2021).

Diabetic macular edema is defined as retinal thickening or the presence of hard exudates involving the macula due to increased vascular permeability in diabetic retinopathy (Parveen et al., 2021; Raman et al., 2025). Structural alterations of retinal capillaries and breakdown of the blood retinal barrier promote fluid accumulation within the macula and progressive visual deterioration if untreated (Rasoulinejad et al., 2015).

Globally, the prevalence of diabetic macular edema is approximately 7.6% (Syndrome et al., 2020), while higher prevalence rates have been reported in Tanzania at 16.1% (Cleland et al., 2016) and 15.3% in Turkey (Acan et al., 2018). The burden of DME is expected to increase substantially as diabetes prevalence is projected to rise by more than 50% worldwide between 2000 and 2030 (Gallagher and Laouri, 2010).

Fundoscopically, DME manifests as focal or diffuse retinal thickening with lipid-rich hard exudates derived from plasma lipoproteins leaking from microaneurysms. Following fluid resorption, lipid deposition commonly occurs within the internal and external plexiform layers and occasionally beneath the neurosensory retina (Rasoulinejad et al., 2015). Untreated disease may result in irreversible visual loss, although laser photocoagulation, intravitreal corticosteroids, and anti-vascular endothelial growth factor therapies have demonstrated significant benefit (Gallagher and Laouri, 2010).

Several epidemiological studies have identified prolonged diabetes duration, poor glycemic control, hypertension, and abnormal serum lipid levels as important risk factors for DME (Agroiya et al., 2013; Benarous et al., 2011). Dyslipidemia is characterized by elevated triglycerides, low-density lipoprotein cholesterol, and total cholesterol levels or reduced high-density lipoprotein cholesterol (Elagamy et al., 2019; Ahmed et al., 2021).

Elevated serum lipid levels contribute to endothelial dysfunction and disruption of the blood retinal barrier, promoting lipid and lipoprotein leakage into retinal tissues and subsequent formation of macular edema (Cardoza et al., 2021). Considering inconsistencies in global findings and limited available local evidence, this study aims to investigate the association between serum lipid abnormalities and diabetic macular edema.

## 2. METHODOLOGY

A hospital based cross-sectional study aimed to evaluate dyslipidemia as a predictor for macula edema in type 2 diabetic patients attending Kilimanjaro Christian Medical Centre (KCMC) from 15/08/2023 to 14/08/2024.

The study enrolled a total of 296 patients with Type 2 diabetes mellitus who were attending the Outpatient clinic at Kilimanjaro Christian Medical Centre (KCMC), using a consecutive non-probability sampling method. Patients were excluded if they had retinal vascular conditions such as retinal vein occlusion, were on lipid-lowering medications, were pregnant, had chronic kidney disease, hemolytic disorders, a history of vitreoretinal surgery, media opacities obstructing fundus visualization, or a prior diagnosis of glaucoma.

Macula edema was assessed as the dependent variable, while the independent variables included triglyceride levels, total serum cholesterol, high-density lipoprotein (HDL), low-density lipoprotein (LDL), duration of diabetes, sociodemographic characteristics, blood pressure, HbA1c, presence of proteinuria, serum creatinine, and body mass index (BMI).

Data were collected using a structured form divided into five sections: informed consent, demographic details, physical examination, diabetic retinopathy grading, macular edema assessment, and laboratory investigations. Demographic data included patient identification number, age, gender, residence, diabetes duration, treatment type, visual acuity, and intraocular pressure. Laboratory evaluations covered serum cholesterol, triglycerides, HDL, LDL, HbA1c, serum creatinine, and urinalysis.

Ethical approval was obtained from the Kilimanjaro Christian Medical University College Research and Ethics Review Committee (Ref: PG 89/2023), with institutional clearance granted via the Ophthalmology Department at KCMC. All participants were informed about the study’s purpose, and written consent was obtained. Participant confidentiality and data protection were maintained throughout.

After obtaining informed consent, visual acuity (VA) and intraocular pressure (IOP) were measured using a Snellen chart and Goldmann applanation tonometer, respectively. Blood pressure was assessed using a digital sphygmomanometer. Comprehensive fundus examination was performed using indirect ophthalmoscopy with a 90D Volk lens on a Haag-Streit slit lamp, following pupillary dilation with 0.5% tropicamide and 5% phenylephrine. Based on retinal findings hard exudates and other features of diabetic retinopathy were recorded, including hemorrhages, proliferative membranes, and neovascularization. A time-domain OCT device (Carl Zeiss, PRIMUS 200; Ref. 000000-2162-427) was used to measure macular thickness. Central subfield macular thickness (CSMT), defined as the mean retinal thickness within the central 1-mm diameter area of the macula, was used to evaluate macular edema. A CSMT value of ≥ 250 µm was considered the cutoff for macular edema.

There after laboratory investigations were conducted, including serum lipid profiling (total cholesterol, triglycerides, HDL, and LDL) using the COBAS INTEGRA 400 PLUS analyzer. A lipid profile was considered abnormal if HDL was <1.15 mmol/L, LDL >3.37 mmol/L, triglycerides >2.26 mmol/L, or total cholesterol >5.2 mmol/L. Additional tests included serum creatinine, glycated hemoglobin (HbA1c), and urinalysis.

Data were analyzed using STATA version 17 (Stata Corp LLC, College Station, TX, USA). Prior to analysis, data cleaning procedures including encoding, labeling, recoding, and variable definition were performed to ensure consistency. Categorical variables were presented as frequencies and percentages, while numerical data were summarized using means with standard deviations or medians with interquartile ranges. Chi-square (χ2) and Fisher’s exact tests were utilized to assess the proportion of central subfield thickness among participant characteristics. A P-value below 0.05 was deemed as indicative of significant variances.

A modifies Poisson regression model was used to assess factors associated with central subfield thickness. Univariate models were first fitted to obtain crude prevalence ratio for factors associated with central subfield thickness. Variables with P-value of less than 0.05 and those of clinical importance in this study, were taken in multivariable models to obtain adjusted prevalence ratio for factors associated with central subfield thickness. Variables with P-value less than 0.05 in multivariable analysis, were considered statistically significant associated with the study outcomes.

## 3. RESULTS

The study included 296 participants with a mean age of 62 years, of whom 55% were female and 45% male. Nearly all participants (about 97%) had a diabetes duration exceeding five years. Systolic hypertension was present in more than 60% of the cohort, while approximately half (51%) were either overweight or obese.

(**Table 1**).

**Table 1:** Social demographic and clinical characteristics of the study participants (N=296)

| **Variable** | **Frequency** | **Percentage** |
| --- | --- | --- |
| **Age** |  |  |
| *Mean (SD)* | 62.4 ($\pm 8.77$) | |
| **Sex** |  |  |
| Male | 132 | 44.6 |
| Female | 164 | **55.4** |
| **Residence** |  |  |
| Urban | 258 | **87.2** |
| Rural | 38 | 12.8 |
| **Duration of DM** |  |  |
| $\leq 5$ Years | 8 | 2.7 |
| $> 5$ Years | 288 | **97.3** |
| *Mean (SD)* | 11 ($\pm 3.53$) | |
| **Type of medicine use** |  |  |
| Oral | 293 | **99** |
| Injectable | 3 | 1 |
| **Intraocular pressure of Right eye** |  |  |
| Normal | 288 | **97.3** |
| High | 8 | 2.7 |
| **Intraocular pressure of Left eye** |  |  |
| Normal | 284 | **95.9** |
| High | 12 | 4.1 |
| **Systolic blood pressure** |  |  |
| $< 140$ | 115 | 38.9 |
| $\geq 140$ | 181 | **61.1** |
| **Diastolic blood pressure** |  |  |
| $< 90$ | 192 | 64.9 |
| $\geq 90$ | 104 | **35.1** |
| **BMI** |  |  |
| Underweight | 4 | 1.4 |
| Normal | 142 | 48 |
| Overweight | 112 | **37.8** |
| Obese | 38 | 12.8 |

Laboratory findings showed that abnormal lipid levels were common: 49% had high total cholesterol, 44% elevated triglycerides, 36% raised LDL, and 39% low HDL. Glycemic control was also poor, with 99% of participants having HbA1c above 6%

(**Table 2**).

**Table 2:** Participants laboratory findings (N=296)

| **Variable** | **Frequency** | **Percentage** |
| --- | --- | --- |
| **Serum creatinine** |  |  |
| *Median (IQR)* | *96.5 (73-140)* |  |
| **Serum cholesterol** |  |  |
| Normal | 152 | 51.4 |
| High cholesterol | 144 | **48.6** |
| *Mean, SD* | *5.4 (±1.47)* |  |
| **Serum triglyceride** |  |  |
| Normal | 167 | 56.4 |
| High triglyceride | 129 | **43.6** |
| *Mean, SD* | *2.1 (±1.35)* |  |
| **Serum LDL** |  |  |
| Normal | 189 | 63.9 |
| High LDL | 107 | **36.1** |
| *Mean, SD* | *2.8 (±1.15)* |  |
| **Serum HDL** |  |  |
| Normal | 181 | 61.1 |
| Low HDL | 115 | **38.9** |
| *Mean, SD* | *1.4 (±0.75)* |  |
| **Urine protein** |  |  |
| Negative | 166 | 56.1 |
| Mild | 31 | 10.5 |
| Moderate | 45 | **15.2** |
| Severe | 30 | 10.1 |
| Trace | 24 | 8.1 |
| **HBA1C (%)** |  |  |
| ≤ 5.81 | 3 | 1 |
| > 5.81 | 293 | **99** |
| *Mean, SD* | *10.3 (±4.7)* |  |

On looking for the factors associated with Macula edema, the proportion of patients with macular edema was found to be 56.4% (167 out of 296), Patients with elevated serum cholesterol levels were found to have a 26% higher likelihood of developing macular edema compared to those with normal serum cholesterol levels (CPR=1.26; 95% CI:1.03-1.55; P-value= 0.023). Similarly, patients with high serum triglyceride levels had a 55% increased risk of macular edema (CPR=1.55; 95% CI:1.2-1.89; P-value < 0.001) compared to individuals with normal serum triglyceride levels.

Furthermore, patients with elevated LDL levels were 27% more likely to develop macular edema compared to those with normal LDL levels (CPR=1.27; 95% CI:1.05-1.55; P-value < 0.001). Lastly, individuals with abnormal urine for protein levels had a 21% higher likelihood of macular edema compared to those with negative or trace levels (CPR=1.21; 95%CI:1.02-1.44; P-value= 0.028).

In the adjusted analysis, patients exhibiting elevated serum triglyceride levels had a 40% higher likelihood of developing macular edema (APR=1.40; 95% CI:1.13-1.74; P-value= 0.002) in comparison to patients with regular serum triglyceride levels. **(Table 3)**

**Table 3:** Factors associated with Macula edema among type 2 diabetic patients (N-296).

| **Variable** | **CPR (95%CI)** | **P-value** | **APR (95%CI)** | **P-value** |
| --- | --- | --- | --- | --- |
| **Sex** |  |  |  |  |
| Male | Ref |  |  |  |
| Female | 0.97 (0.79-1.19) | 0.821 |  |  |
| Age | 1.004 (0.99-1.07) | 0.420 |  |  |
| **Duration of diabetic mellitus** | 1.01 (0.99-1.02) | 1.158 |  |  |
| **BMI** | 1.11 (0.98-1.28) | 0.097 |  |  |
| **Serum creatinine** | 1.0001 (0.99-1.001) | 0.880 |  |  |
| **Serum cholesterol** |  |  |  |  |
| Normal | Ref |  | Ref |  |
| High cholesterol | 1.26 (1.03-1.55) | **0.023** | 1.09 (0.89-1.38) | 0.343 |
| **Serum triglycerides** |  |  |  |  |
| Normal | Ref |  | Ref |  |
| High triglyceride | 1.55 (1.27-1.89) | **< 0.001** | 1.40 (1.13-1.74) | **0.002** |
| **Serum HDL** |  |  |  |  |
| Low HDL | 1.08 (0.88-1.32) | 0.449 | 1.04 (0.85-1.26) | 0.736 |
| Normal | Ref |  | Ref |  |
| **Serum LDL** |  |  |  |  |
| Normal | Ref |  | Ref |  |
| High LDL | 1.27 (1.05-1.55) | **0.015** | 1.17 (0.95-1.44) | 0.138 |
| **Urinalysis protein** |  |  |  |  |
| Normal | Ref |  | Ref |  |
| Abnormal | 1.21 (1.02-1.44) | **0.028** | 1.14 (0.94-1.39) | 0.188 |
| **HBA1C** | 0.99(0.97-1.02) | 0.999 |  |  |

## 4. DISCUSSION

Abnormal lipid profiles were highly prevalent, with nearly half of participants demonstrating elevated total cholesterol, highlighting a substantial burden of dyslipidemia in the study population. This pattern may reflect sedentary lifestyles, unhealthy dietary habits, and limited access to healthcare services. Similar observations were reported in Kishanganj, India, where elevated cholesterol, triglycerides, and low HDL were observed in 34%, 26%, and 26% of patients, respectively (Mandal, Kumari, and Mukherjee, 2015). Higher prevalence’s were documented in Bangladesh 52% for cholesterol, 58% for triglycerides, 50% for low HDL, and 74% for LDL abnormalities (Ahmmed et al., 2021) while Rwanda reported a lower prevalence of 19% (Niyibizi et al., 2018). These variations may reflect genetic influences, including APOE, PCSK9, and LPL gene differences, alongside environmental and lifestyle factors across populations.

This study identified a significant proportion of macular edema at 56.4% (167 out of 296), possibly due to advanced stages of diabetic retinopathy and longer durations of diabetes among our participants. In contrast, a study conducted in Mexico (Graue-Hernandez *et al*., 2020) reported a much lower proportion of 6.6% for macular edema. The variation can be attributed to the shorter duration of diabetes (less than 5 years) among the participants in the Mexican study.

In finding the relationship between dyslipidemia and macular edema, the study highlights that elevated serum cholesterol, triglycerides, and LDL levels were significantly associated with macular edema. Even after adjustment, serum triglycerides remained significantly associated with 40% higher risk on the occurrence of macular edema. This could be attributed to the fact that elevated lipid levels in retinal capillaries promote the production of pro-inflammatory cytokines, leading to chronic inflammation and the formation of reactive oxygen species (ROS) which damages the endothelial cells lining retinal blood vessels, contributing to increased capillary leakage and consequently macular edema formation.

These findings were consistent with the study by (Kolli, Endreddy and Reddy, 2015) in India but differed from the study by (Cetin *et al*., 2013) in Turkey, which found no link between dyslipidemia and macular edema. The variations may be due to the fact that most patients in the Turkish study were normotensive as compared to our study, which also incorporates the concept of metabolic syndrome in the progression of diabetic retinopathy and macula edema. Furthermore, our study involved a larger sample size compared to the Turkish study.

A key limitation in this study is that being hospital-based, the study’s sample may not represent the broader population of type 2 diabetic patients, potentially overestimating the prevalence of macula edema and dyslipidemia due to a higher concentration of patients with more advanced disease or complications seeking treatment at hospitals.

## 5. CONCLUSION

The study demonstrated a significant association between dyslipidemia and macular edema. The findings highlight the importance of early intervention and lifestyle modification in the management of diabetes and its related complications. They also underscore the need for strengthened screening programs and improved patient education to help reduce the occurrence and progression of macular edema.

## Data Availability

All data produced in the present study are available upon reasonable request to the authors

## 6. RECOMMENDATIONS

Future studies should emphasize large, methodologically robust investigations, including case control designs, to more clearly determine the causal link between dyslipidemia and the progression of macula edema. In clinical practice, integrating routine lipid assessment, patient counseling, and lifestyle modification into diabetes management is essential. Enhanced multidisciplinary collaboration, together with supportive national health policies, is equally important to promote early identification, optimal lipid control, and prevention of vision threatening diabetic complications.

## DISCLAIMER (ARTIFICIAL INTELLIGENCE)

The authors hereby declare that no artificial intelligence tools, including large language models or text-to-image generators, were used at any stage of the preparation, writing, or editing of this manuscript. The work presented represents the authors’ independent and original research.

## ACKNOWLEDGEMENT

I thank Dr. William Makupa, Dr. Andrew Makupa and Frank Sandi for their invaluable mentorship and guidance.

I sincerely appreciate Kalvin Rwegoshora for his support in data analysis and biostatistics. Grateful to CBM for financial support and to my family for their constant encouragement.

